# Quantitative MODS-Wayne Assay for Rapid Detection of Pyrazinamide Resistance in *Mycobacterium tuberculosis* from Sputum Samples

**DOI:** 10.1101/2024.01.05.24300793

**Authors:** Emily Toscano-Guerra, Roberto Alcántara, Robert Gilman, Louis Grandjean, Mirko Zimic, Patria Sheen

## Abstract

**Background:** Tuberculosis (TB) remains a significant global health challenge, particularly with the rise of drug-resistant strains, such as Pyrazinamide (PZA)-resistant TB. This resistance hampers treatment effectiveness. Currently, there’s a lack of affordable and precise quantitative tests for detecting PZA resistance, highlighting the need for accessible diagnostic tools. Our study introduces a direct, accurate, and accessible susceptibility test for PZA resistance by quantifying pirazinoic acid (POA).

**Methods:** We analyzed 264 TB-positive samples (MP and TBN) to assess PZA susceptibility by measuring POA production. This was done using the MODS-Wayne qualitative assay and our developed quantitative approach (MODS-WQ). For MODS-WQ, TBN samples were processed in 7H9 media (MODS-WQ 7H9) and MP samples in citrate buffer (MODS-WQ CB). We measured POA levels using spectrophotometry against a calibration curve, with PZA susceptibility determined by a composite standard. We also investigated the correlation between POA levels and pyrazinamidase mutations. Multidrug-resistant samples were assessed using the MODS test, and statistical analyses were conducted.

**Results:** Approximately 23.48% of the samples showed PZA resistance, which included 62.8% of the multidrug-resistant (MDR) samples. The MODS-Wayne assay demonstrated an 87.8% sensitivity and 95.9% specificity for MP samples, and 62.5% sensitivity with 93.7% specificity for TBN samples. The MODS-WQ identified specific cut-off points—123.25 μM for MODS-WQ 7H9 and 664.7 μM for MODS-WQ CB—with corresponding sensitivities of 81.25% and 92.31%, and specificities of 77.22% and 95.93%. Significantly lower POA levels were observed in samples with mutations in the metal binding site compared to mutations on the enzyme’s periphery.

**Conclusions:** The citrate-buffered MODS-WQ demonstrates high sensitivity and specificity in quantifying POA. It offers a significant improvement over qualitative methods, effectively overcoming their subjectivity limitations in PZA resistance detection.

## BACKGROUND

Tuberculosis (TB) remains a major global health concern, particularly in low- and mid-income countries where it has reached pandemic proportions. The emergence of drug-resistant TB strains poses a significant challenge to eradication efforts [1]. Pyrazinamide (PZA), a first-line anti-TB drug, is pivotal in shortening treatment duration to six months for both drug-sensitive and multidrug-resistant (MDR) TB cases [2]. PZA acts as a prodrug, which is converted into its active form, pyrazinoic acid (POA), by the enzyme pyrazinamidase (PZase). The PZase enzyme is encoded by the pncA gene, playing a crucial role in the drug’s efficacy [3].

Resistance to Pyrazinamide (PZAr) significantly worsens TB treatment outcomes and increases mortality rates [4]. It’s estimated that PZAr occurs in about 16.2% of all TB cases globally [5] and constitutes approximately 60% of multidrug-resistant (MDR) TB cases. The lack of a standardized and reliable method for routinely detecting PZAr presents a major obstacle in TB management strategies [6].

Routine testing for PZA resistance (PZAr) is often not performed due to various technical challenges [7]–[9]. Existing phenotypic culture-based drug susceptibility testing (pDST) methods, such as the BACTEC MGIT 960 system [10], the Wayne assay [11], the Nitrate reductase assay [12], and the Microscopic Observation Drug Susceptibility (MODS) assay [13], face several limitations. These include difficulties in maintaining the precise inoculum amount and pH level (5.5-6.5) necessary for PZA activity, the risk of contamination with other bacterial strains leading to an error rate of 10-15% [7], and prolonged detection times [8]. Additionally, the Bactec MGIT 960 system, despite its high cost, is known to produce a significant rate of false resistance results [9].

The Wayne assay, which assesses the enzymatic activity of pyrazinamidase (PZAse), is an economical colorimetric test that identifies pyrazinoic acid (POA) as a marker of susceptibility using ferrous ammonium sulfate (SAF) [11]. Despite its cost-effectiveness, this assay has limitations: it is qualitative and subjective, and it necessitates extended incubation periods until bacterial growth is detectable [8].

Genotypic drug susceptibility testing (gDST) methods, such as Sanger sequencing and whole-genome sequencing, are employed to identify specific genetic mutations in the pncA gene linked to PZA resistance (PZAr) [14], [15]. These methods can also detect mutations in other genes, including *rpsA*, *panD*, and *clpC1* [16]–[19]. gDSTs are generally more sensitive and specific than phenotypic methods, with the majority (70-97%) of PZA-resistant clinical isolates showing genetic variations in the pncA gene’s promoter region or coding sequence [20], [21]. However, these techniques require significant technical expertise and are costly, making them less accessible in low- and mid-income countries where TB is endemic [22]. Additionally, the variability in mutation patterns linked to PZAr can differ regionally, affecting the representativeness of these tests [23].

The Microscopic Observation Drug Susceptibility (MODS) assay is an effective, low-cost, and simple method for diagnosing TB and MDR-TB directly from sputum samples, with a short turnaround time of 5 to 21 days. However, its application for Pyrazinamide (PZA) testing has been limited. To address this, Alcántara et al. [24] developed the MODS-Wayne method, combining the MODS assay with the Wayne test. This adaptation is based on detecting pyrazinoic acid (POA), the hydrolyzed product of PZA expelled into the extracellular environment, via a colorimetric reaction with ferrous ammonium sulfate (SAF) [11]. The MODS-Wayne assay demonstrated high sensitivity and specificity (92.7% and 99.3%, respectively). Nonetheless, its subjective nature may affect the reproducibility of results.

In this study, we aim to refine the MODS-Wayne test by introducing a quantitative approach (MODS-WQ) using sputum samples from TB patients. This new approach seeks to reduce diagnosis time, eliminate the need for pre-isolation in solid cultures, and move beyond qualitative judgments.

## MATERIALS AND METHODS

### Samples and Study design

264 TB-positive samples from two studies conducting during 2015-2016 (coded as MP) and 2017-2018 (coded as TBN), obtained from patients enrolled in the National TB Strategy treatment program at the Hospital Nacional Dos de Mayo, Lima, Peru, and from the Regional Tuberculosis Reference Laboratory, Callao, Lima, Peru. Only basic data (age, sex, treatment) was recorded to the sample anonymity. This work was approved by the Institutional Committee on Research Ethics (CIEI) of the Universidad Peruana Cayetano Heredia (Code 331-34-21). For the quantitative MODS-WQ test, two variants were performed and contrasted with the qualitative version MODS-Wayne. One variant (MP n=169), were carried out in citrate buffer (pH 7.0) matrix, named as MODS-WQ BC, whereas the other (TBN n=95) was coded as TBN, which were performed in the same media culture 7H9 (pH ̴ 6.8), named as MODS-WQ 7H9 (**Figure 1**).

**Figure 1.**
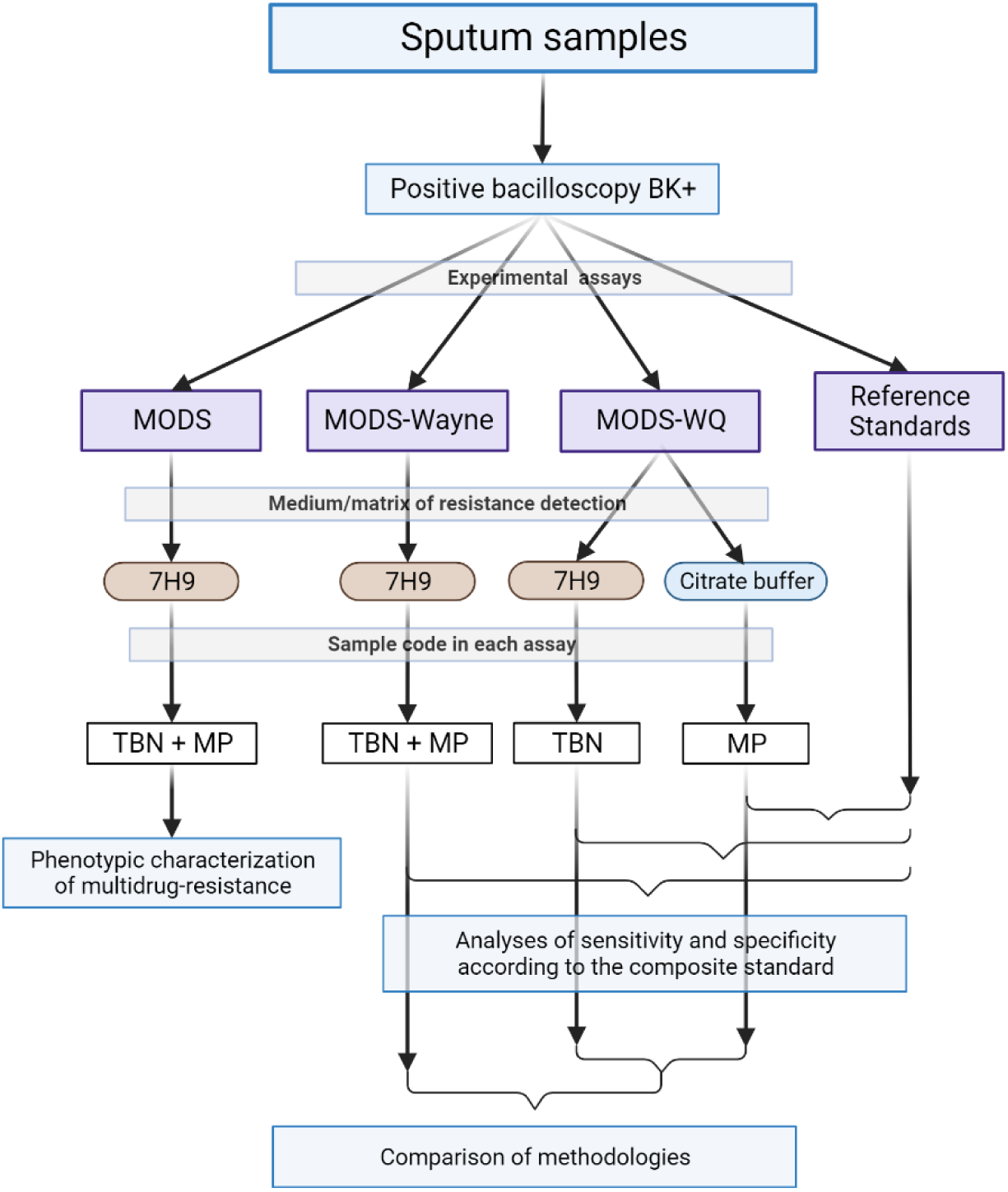
Study design. Sputum samples were collected from two studies: MP and TBN. Only samples positive for bacilloscopy were included. Samples were used in four experimental assays: MODS (MDR characterization), MODS-Wayne (qualitative assay for PZA resistance), MODS-WQ (quantitative variant) and three reference standards. The diagnostic value for MODS-Wayne and MODS-WQ was calculated using the standard composite.

### Multidrug resistant test: rifampicin (RIF) and isoniazid (INH) resistance

All sputum samples were previously decontaminated according to the protocol by Kent and Kubica [25] and seeded in 7H9 Middlebrook (**See Supplementary methods**). MODS test was carried out using the protocol implemented in our laboratory [26]. Samples were incubated with INH at final concentration of 0.4 µg/ml or RIF at 1.0 µg/ml at 37°C for a maximum of 21 days fallowed by evaluation using an inverted microscope.

### MODS-Wayne qualitative assay for PZA susceptibility

This test was conducted simultaneously with the MODS-MDR assay (**Supp. Figure S1A**). Samples were allocated into two wells: Control (PZA-C) and treatment (PZA-Wayne), then incubated at 37°C. Upon observing growth in the PZA-C well, the date was recorded, and incubation continued for an additional three days. Subsequently, PZA (800 μg/mL) was introduced into the PZA-Wayne well and further incubated for three days. Next, 100 μL of 10% SAF was added and incubated for five minutes, with immediate color registration. A red color indicated sensitivity, while the absence of color denoted resistance. Due to variations in observed coloration among samples, an intensity scale from 0 to 3 was established, where 0 represented resistance and 3 signified high sensitivity (**Supp. Figure S1C-D**). For more details **see supplementary methods**.

### MODS-WQ assay for PZA susceptibility

This assay included two variants: MODS-WQ BC (10 mM citrate pH 7.0) and MODS-WQ 7H9 (pH ∼6.8), performed in separate 24-well plates (**Supp. Figure S1B**). Samples were incubated as in MODS-Wayne assay. Following a similar incubation process to the MODS-Wayne assay, upon observing positive growth in the PZA-C well on the third day, samples underwent variant-specific treatments. For MODS-WQ 7H9, samples were completed to 1 mL by adding 100 μL of 7H9 media to the PZA-C well and 100 μL of PZA (800 μg/mL final concentration) to the PZA-WQ well, followed by an additional three-day incubation. Conversely, in the MODS-WQ CB procedure, samples were centrifuged, the resulting pellet resuspended in 900 μL of citrate buffer and returned to a 24-well plate. Then, 100 μL of citrate buffer was added to the PZA-C well and 100 μL of PZA (800 μg/mL final concentration) to the PZA-WQ well, also incubated for three days.

Post-incubation, 100 μL of 10% SAF was added and incubated for 5 minutes. Subsequently, 500 μL was transferred to cryotubes and stored at −80°C for subsequent analysis. During the analysis, triplicates of samples (100 μL) were transferred to a 96-well plate and measured at a 450nm wavelength using a spectrophotometer. Notably, MODS-WQ 7H9 samples were centrifuged (13000 rpm, 2 minutes) and supernatants were analyzed. Absorbance values were then interpolated against standard POA curves, ranging from 31.25 µM to 4000 µM (Table S1). Calibration curves with 7H9 matrix underwent also previous centrifugation (13000 rpm, 2 minutes), with subsequent measurement of supernatants. Mixture triplicates (100 μL) were analyzed on a spectrophotometer at a 450nm wavelength to determine absorbance values, for calculating linear regression on standard curves). For more details **see supplementary methods**.

### Composite reference standard for PZA susceptibility determination

In the absence of a singular standard test, we worked with a composite standard, based on three tests: (i) the BACTEC MGIT960 (ii) conventional Wayne test, and (iii) sequencing of the *pncA* gene (**Supp. Figure S2A**). Each sample was coded as 0 (sensitive) or 1 (resistant) based on its response to these three standard tests. Samples were then categorized as resistant (summing up to 2 or 3) or sensitive (summing up to 0 or 1) within the composite standard (**Supp. Figure S2B**). Mutations were classified according to WHO catalog[27] and the SuspectPZA webtool: https://biosig.lab.uq.edu.au/suspect_pza/. Both MODS-Wayne and MODS-WQ were evaluated against this composite standard. More details in **supplementary methods**.

### Evaluation of PZase mutations effect on POA production

To assess the impact of enzyme mutations on pyrazinoic acid (POA) production, POA values (µM) were plotted per sample according to mutation sites, categorized as Active Catalytic Site (ACS), Metal Binding Site (MBS), PZA Binding Site (PBS), Enzymatic Core (EC), and the Enzyme’s Periphery (PER). POA production was compared to that of the wild-type enzyme to evaluate alterations caused by mutations.

### Statistical analysis

Continuous variables were expressed as median ± IC95% or median and interquartile range (IQR). Sample comparisons were made using non-parametric Mann-Whitney test. The sensitivity and specificity of qualitative MODS-Wayne was estimated using a 2-by-2 contingency table. The predictive value of MODS-WQ assay was evaluated with the logistic regression model to calculate the Receiver Operating Characteristic (ROC) curve, sensitivity and specificity of the test. Agreement between assays and the composite standard was estimated using the Kappa index. Statistical tests were performed with STATA 14 and GraphPad Prism 9.0.2. The analysis of mutations was performed with the Pymol 2.5.4 educational license software.

## RESULTS

### Study population and MDR characterization

Patient characterization is shown in **Table 1**. The MP and TBN groups exhibited no significant differences in age and gender distribution. In MP group only 11.00% of patients had received prior treatment, while in the TBN group, 67.00% had received it. Among the 264 collected samples, 78 (29.54%) were identified as MDR samples. Within the non-MDR samples (n = 186, 70.45%), 26 demonstrated monoresistance to INH (9.8%), 15 displayed monoresistance to RIF (5.7%), and 145 samples exhibited pansusceptibility (55.30%).

**Table 1.**
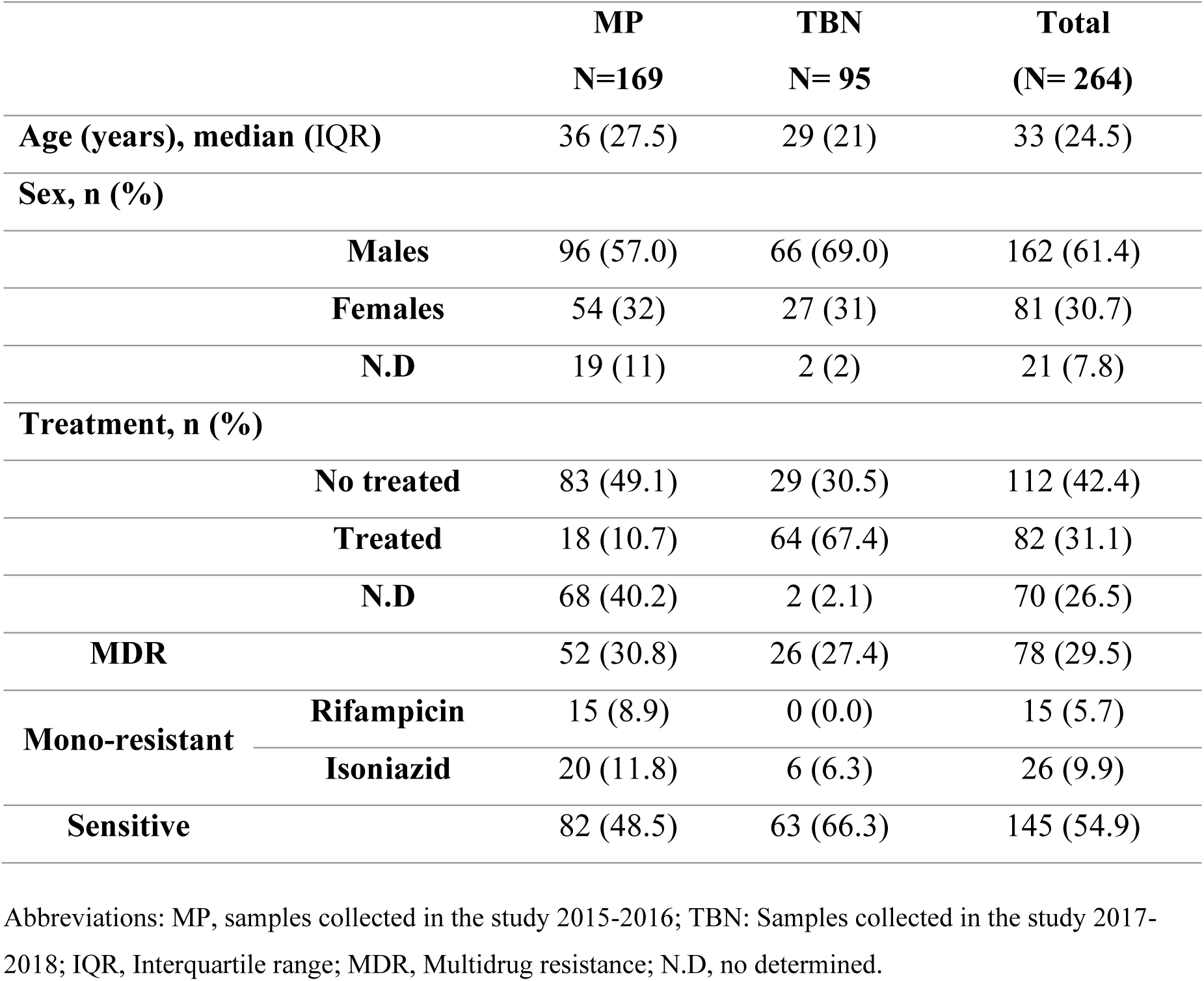
Baseline characteristics of the samples.

### Composite reference standard tests for PZA susceptibility determination

In the TBN group (n = 95), MGIT-PZA identified 80 isolates (84.2%) as sensitive and 15 (15.8%) as resistant. Conversely, in the MP group (n = 169), 122 isolates (72.2%) were sensitive, while 47 (27.8%) were resistant. The conventional Wayne test showed sensitivity in 83 (87.4%) isolates and resistance in 12 (12.6%) within the TBN group; for the MP group, 137 (81.1%) were sensitive, and 32 (18.9%) were resistant. The *pncA* sequencing revealed 187 wild-type isolates (TBN: n = 71, MP: n = 116) and 69 isolates with resistant mutations, (undetermined in 8 isolates). Among the mutations (n = 19), 16 were point mutations, 2 deletions, and 2 insertions (Table 3). Notably, 1 (1.5%) affected the promoter site, 1 (1.5%) affected the catalytic site, 4 (5.8%) targeted the metal binding site, 5 (7.3%) were located in the periphery, and 6 (8.7%) affected the enzyme core (**Table 2**). Following the three tests’ susceptibility results, a composite susceptibility was calculated for each isolate (**Table 3**).

**Table 2.**
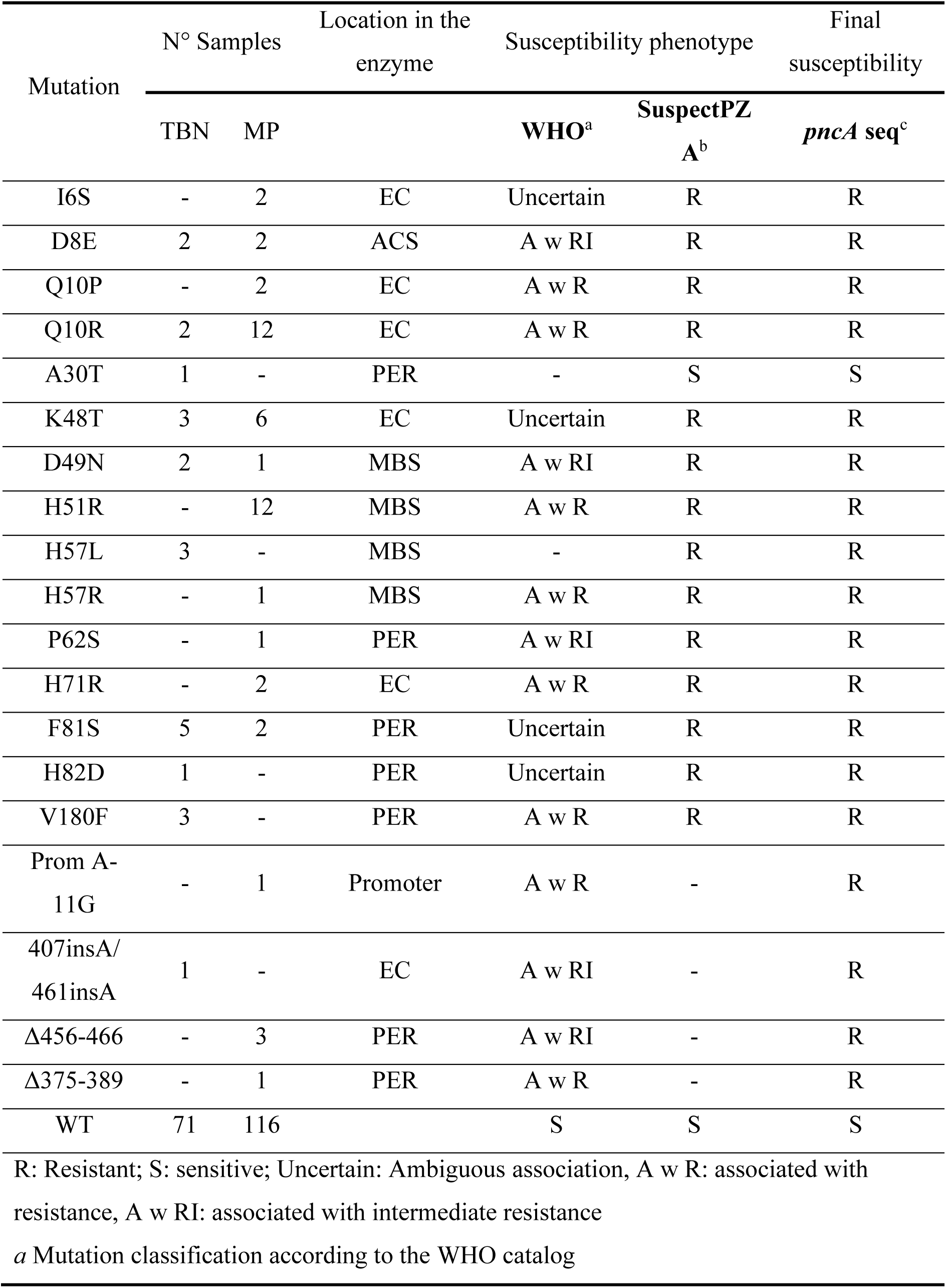
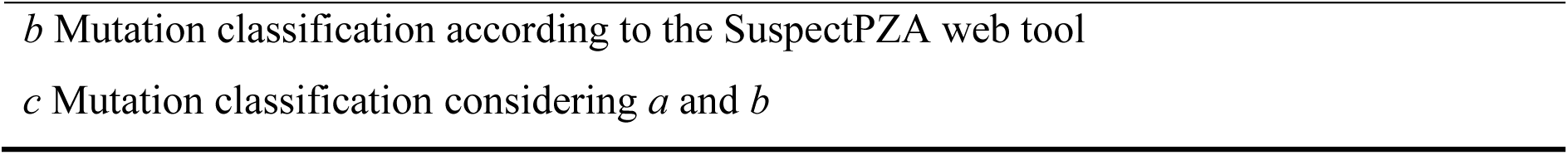
Categorization and characterization of the mutations found in the study.

| Mutation | N° Samples |  | Location in the enzyme | Susceptibility phenotype |  | Final susceptibility |
| --- | --- | --- | --- | --- | --- | --- |
|  | TBN | MP |  | WHO<sup>a</sup> | SuspectPZ A<sup>b</sup> | *pncA* seq<sup>c</sup> |
| I6S | - | 2 | EC | Uncertain | R | R |
| D8E | 2 | 2 | ACS | A w RI | R | R |
| Q10P | - | 2 | EC | A w R | R | R |
| Q10R | 2 | 12 | EC | A w R | R | R |
| A30T | 1 | - | PER | - | S | S |
| K48T | 3 | 6 | EC | Uncertain | R | R |
| D49N | 2 | 1 | MBS | A w RI | R | R |
| H51R | - | 12 | MBS | A w R | R | R |
| H57L | 3 | - | MBS | - | R | R |
| H57R | - | 1 | MBS | A w R | R | R |
| P62S | - | 1 | PER | A w RI | R | R |
| H71R | - | 2 | EC | A w R | R | R |
| F81S | 5 | 2 | PER | Uncertain | R | R |
| H82D | 1 | - | PER | Uncertain | R | R |
| V180F | 3 | - | PER | A w R | R | R |
| Prom A-11G | - | 1 | Promoter | A w R | - | R |
| 407insA/<br>461insA | 1 | - | EC | A w RI | - | R |
| Δ456-466 | - | 3 | PER | A w RI | - | R |
| Δ375-389 | - | 1 | PER | A w R | - | R |
| WT | 71 | 116 |  | S | S | S |

R: Resistant; S: sensitive; Uncertain: Ambiguous association, A w R: associated with resistance, A w RI: associated with intermediate resistance
<sup>a</sup> Mutation classification according to the WHO catalog
*b* Mutation classification according to the SuspectPZA web tool
*c* Mutation classification considering *a* and *b*

**Table 3.** PZA susceptibility results according to composite standard.

| Table 3. PZA susceptibility results according to composite standard |  |  |  |  |  |  |
| --- | --- | --- | --- | --- | --- | --- |
| Phenotype | Sum value | Standard composite |  |  |  | Total (264) |
|  |  | MP (169) |  | TBN (95) |  |  |
|  |  | N (%) | N Subtotal | N (%) | N Subtotal |  |
| Resistant | 0 (%) | 30 (17.8) | 46 (27.2) | 8 (8.4) | 16 (16.8) | 62 (23.5) |
|  | 1 (%) | 16 (9.5) |  | 8 (8.4) |  |  |
| Sensitive | 2 (%) | 10 (5.9) | 123 (72.8) | 10 (10.5) | 79 (83.2) | 202 (76.5) |
|  | 3 (%) | 113 (66.9) |  | 69 (72.6) |  |  |

### MODS-Wayne qualitative assay performance

In the MP group, based on the observed color scale, 24.26% of samples were classified as PZA-resistant, while 75.74% were deemed sensitive. Conversely, in the TBN group, 15.79% were identified as resistant, and 84.21% were labeled as sensitive to PZA (Table S4).

For assessing MODS-Wayne’s sensitivity and specificity, the obtained proportions were compared to those derived from the composite standard. The results indicated an 87.8% sensitivity (95% CI: 0.7446 - 0.9468, Wilson-Brown test) and 95.9% specificity (95% CI: 0.9084 - 0.9825), accompanied by a kappa index of 0.83 (p=0.001) (**Table 4**).

**Table 4.** Comparative performance of each MODS-Wayne and MODS-WQ methodology with their variants in CB and 7H9.

| Method/Sample |  | Ratio | Sensitivity [95% CI] | Ratio | Specificity [95% CI] | PPV | NPV | Ki |
| --- | --- | --- | --- | --- | --- | --- | --- | --- |
| **MODS-Wayne** | **MP** | **36/41** | **0.878 [0.745-0.947]** | **118/123** | **0.959 [0.901-0.983]** | **0.88** | **0.95** | **0.83** |
|  | TBN | 10/16 | 0.625 [0.387-0.815] | 74/79 | 0.937 [0.860-0.973] | 0.67 | 0.93 | 0.57 |
| **MODS-WQ** | **MP-BC <665 µM** | **38/41** | **0.923 [0.797-0.974]** | **118/123** | **0.959 [0.908-0.983]** | **0.88** | **0.98** | **0.87** |
|  | TBN-7H9 <127 µM | 13/16 | 0.813 [0.569-0.934] | 61/79 | 0.772 [0.668-0.851] | 0.42 | 0.95 | 0.43 |

Measures of accuracy (sensitivity, specificity, positive predictive value PPV, negative predictive value NPV) and confidence intervals were observed for each type of test evaluated.

### MODS-WQ assay performance

The samples were categorized into sensitive and resistant groups based on the composite standard. In MODS-WQ 7H9 samples, the median POA production according calibration curve (**Figure 2A**) for the sensitive group was 274 μM (IQR = 373.3 μM), contrasting with 92 μM (IQR = 77.95 μM) for the resistant group (**Figure 2B**). The Mann-Whitney test yielded a significant p-value (<0.0001) although no pronounced discrimination was observed. In MODS-WQ BC samples, the median concentration for the sensitive group was 2105 μM (IQR = 1672 μM), whereas for the resistant group, it was 258 μM (IQR = 1383.3 μM) (**Figure 2B**), revealing a strong distinction with a p-value of <0.0001.

**Figure 2.**
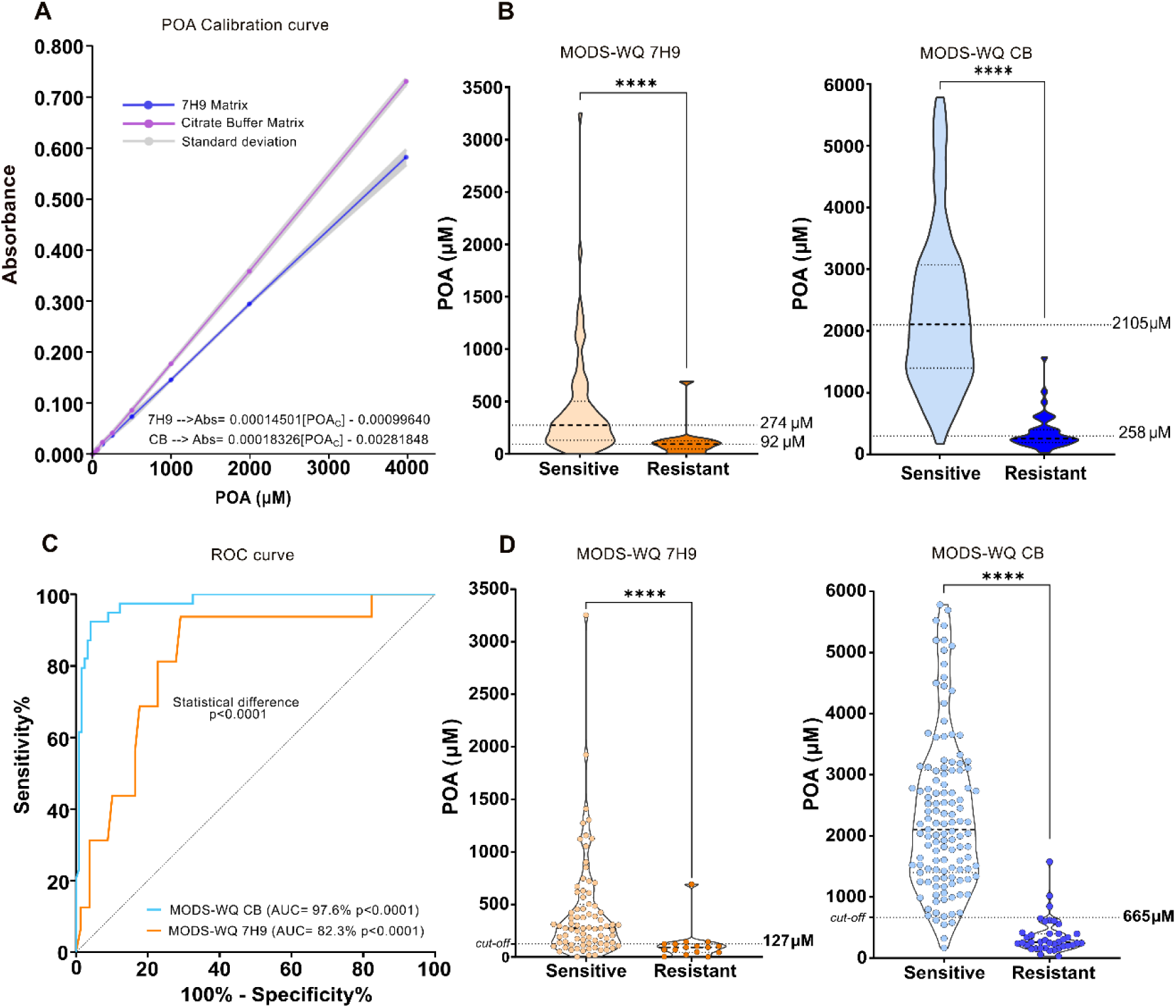
Performance of the MODS-WQ variants. **A)** Standard curves of pyrazinoic acid dissolved in two types of matrices. Curves are result of 3 repetitions and their respective standard deviations. The equation for calculating POA concentration (POAc) is shown. **B)** Violin graph of the classification of TBN and CB samples according to the composite standard test and POA values in μM obtained with the MODS-WQ 7H9 and CB respectively. The medians of POA are shown for each group. **C)** ROC curves and p-values obtained with the two variants MODS-WQ BC and MODS-WQ 7H9, statistical differences are observed between both variants. **D)** Violin graph of classifieds according to MODS-WQ 7H9 and MODS-WQ BC showing the cut-off point for each variant. *p<0.05, ** p<0.01, *** p<0.001, **** p<0.0001

ROC curve analysis displayed good performance for both variants. However, the area under the curve (AUC) for MODS-WQ BC was 97.57% (95% CI: 95.30% – 99.84%), significantly surpassing (p-value= 0.0037) MODS-WQ 7H9 (AUC = 82.28%, 95% CI: 71.28% - 93.28%)

(**Figure 2C**), indicating superior discrimination for MODS-WQ CB. The optimal cut-off points associated with the ROC curves were 126.70 μM for MODS-WQ 7H9, demonstrating a sensitivity of 81.25% (95% CI: 56.99% - 93.41%) and a specificity of 77.22% (95% CI: 66.83% - 85.07%) (**Table 4**-**Figure 2D**). Conversely, for MODS-WQ CB, the optimal cut-off was 664.7 µM, showcasing a sensitivity of 92.31% (95% CI: 79.68% – 97.35%) and a specificity of 95.93% (95% CI: 90.84% - 98.25%) (**Table 4-Figure 2D**). The kappa index resulted in 0.83 (p-value=0.001) for MODS-WQ 7H9 and 0.87 (p-value=0.001) for MODS-WQ BC.

### Relationship between POA production and PZase mutations

The advantage of determining resistance quantifying POA production rather than relying solely on qualitative determination (MODS-Wayne) lies in establishing resistance levels and correlating them with enzymatic activity. Mutations in *pncA* gene can impact in the enzyme activity, so that, we assessed POA production in isolates with mutations across different enzyme regions (**Figure 3A**), excluding deletions and mutations at the promoter site. The **Figure 3B**, shows that mutations within the EC (I6S, Q10P, Q10R, K48T, and H71R) exhibited a median POA production of 252 μM. Mutations in the MBS (D49N, H51R, H57R) caused a median of 266.4 μM. A single mutation in the ACS (D8E) detected in two isolates demonstrated widely varying POA values of 59 μM and 847 μM (median: 453 μM). In contrast, mutations in PER (P62S, F81S) revealed a median of 644.3 μM, close to the cut-off. The median POA production in wild-type isolates was 2195 μM, indicating that mutations in the EC and MBS regions decreased POA levels by approximately 88.5% and 88.0%, respectively. Meanwhile, mutations in PER decreased POA production by approximately 70.6%.

**Figure 3.**
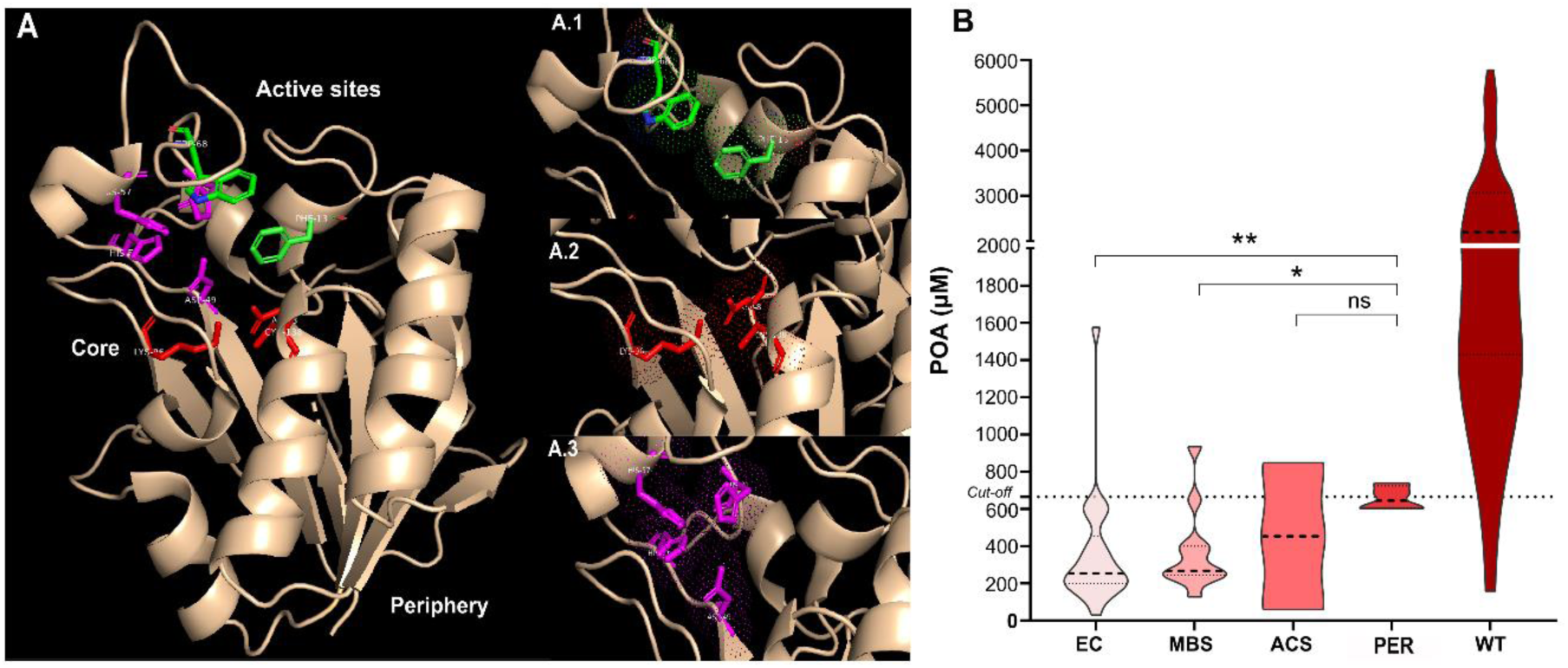
Effect of pyrazinamidase mutations on POA production. **A)** 3-D structure of pyrazinamidase, the active sites are depicted in green, violet and red. Enzyme core area near to active sites and the periphery distant to active sites. Green: Binding site for PZA formed by the amino acids Phe13 and Trp68 (A.1) Red: Catalytic triad, composed of Asp8, Lys96, and Cys138 (A.2) Purple: Metal-binding site (Fe+), consisting of Asp49, His51, and His57 (A.3). **B)** Distribution of POA values at each mutation site, as well as in wild type samples. The MODS-WQ cut-off (665 µM) is indicated by a dashed line. EC: Enzymatic Core, N=22; MBS: Metal-Binding Site, N=11; ACS: Active Catalytic Site, N=2; Periphery: N=4, WT: Wild type, N=116. Mann-Whitney test, * p<0.05, ** p<0.01, *** p<0.001, **** p<0.0001, ns: not significant.

### DISCUSSION

Prompt detection of PZAr is crucial for effective TB treatment. Traditional culturing methods, however, face difficulties in maintaining appropriate pH levels and inoculum concentrations. In our study of 264 samples, 78 (29.54%) were identified as multidrug-resistant (MDR). Among these MDR samples, 49 (62.80%) exhibited resistance to PZA, as determined by a composite standard. This finding aligns with other studies reporting PZAr rates of 38% to 65% in MDR TB cases [28]–[30].

The citrate buffer variant of the MODS-Wayne quantitative (MODS-WQ CB) assay demonstrated outstanding efficacy, with a sensitivity of 92.31%, specificity of 95.93%, and a positive predictive value (PPV) of 88%. In contrast, the 7H9 matrix variant showed notably lower performance, with a sensitivity of 81.3%, specificity of 77.2%, and a PPV of 42%.

The spectrophotometric measurement of POA in 7H9 media is complicated by the presence of bovine serum albumin (BSA), which is added to foster mycobacterial growth [31]. BSA exhibits a higher affinity for POA compared to PZA at concentrations greater than 0.4%, significantly raising the POA minimum inhibitory concentration (MIC) [32]. Additionally, BSA tends to interact with various ligands, particularly divalent metallic cations, including copper and zinc. Notably, BSA shows a strong affinity for Fe2+ and Fe3+ ions at neutral pH, particularly near tryptophan residues, similar to lactoferrin [33]–[35]. This affinity for POA and iron could hinder the formation of the SAF-POA complex, leading to variable sensitivity and specificity in our assay results.

The use of the citrate buffer (CB) matrix in our study not only simplified colorimetric observation but also ensured more accurate absorbance detection by minimizing medium-related biases. This contrasts with previous research by Meinzen et al. [36], which quantified POA using similar matrices but relied on samples grown on agar and stored in a culture bank. Our approach, in contrast, involves direct sampling from patient sputum. Meinzen et al. reported sensitivity and specificity rates of 96.0% and 97.4%, respectively, using a PZA concentration of 400 μg/mL, and 90.5% and 94.4% for 800 μg/mL PZA, achieving an area under the curve (AUC) of over 93.0% for both concentrations.

While our study did not match the high sensitivity and specificity levels reported by Meinzen et al., our approach, using direct patient samples, significantly reduced the diagnosis time. On average, we achieved a 13.5-day reduction compared to traditional methods like the Wayne Test (30-45 days) [24] and the MODS-Wayne (mean 19.5 days). Although the performance of MODS-Wayne and our quantitative MODS-WQ were comparable (Kappa indices of 0.83 and 0.87, respectively), it’s crucial to note the variability in MODS-Wayne results across different sample types (MP, TBN, and previous study), mainly due to observer subjectivity. This variability could affect reproducibility [37], [38], an issue potentially resolved by our quantitative MODS-WQ variant. Further studies focusing on reproducibility are essential to validate our findings.

POA production is a key indirect marker of PZase enzymatic activity, which can be influenced by protein mutations [39]–[41]. In our study, mutations within the PZase enzyme were categorized into four areas: ACS, MBS, EC, and PER. We observed that mutations in the EC and MBS regions led to significantly lower POA levels (252.7 µM and 266.4 µM, respectively) compared to those in PER, which produced POA levels near the 665 µM cut-off (p<0.01 and p<0.05, respectively). Supporting our findings, Sheen et al. [39] reported that mutations in MBS (H51R, D49N) resulted in a substantial decrease in enzymatic activity (99%) in terms of POA production, compared to the wild-type H37Rv strain. Similarly, mutations near active sites (core) led to a 72% reduction, while peripheral mutations caused a 47% decrease in enzymatic activity.

Our study aligns with previous research, showing a marked decrease in POA production in samples with mutations in the EC and MBS regions (88%) compared to wild-type samples. In samples with PER mutations, this reduction was slightly lower at 70.6%. It’s important to distinguish our methodology from that of Sheen et al., who directly quantified POA production in purified enzymes in vitro, free from external influences on the mycobacteria’s POA output.

The release of POA is influenced not only by enzymatic efficiency but also by external factors, such as efflux pump activity, which plays a crucial role in POA transport. Zimic et al. [42] highlighted a strong link between POA efflux rate and PZA resistance. Their study showed that resistant isolates have lower efflux rates, while sensitive isolates display higher rates [40], allowing for the prediction of resistance with high sensitivity and specificity.

Enzymatic activity alone accounts for only 27.3% of PZA resistance cases [39], while the POA efflux rate contributes to around 51% [41]. Notably, 70-90% of PZA-resistant strains show mutations in the *pncA* gene and its promoter region [43]–[45]. This suggests that a significant portion of resistant strains may involve other, yet unidentified, mechanisms contributing to PZA resistance.

Mutations in other genes, such as *panD* and *rpsA*, might also play a role in PZA resistance, as could the presence of heteroresistant strains with varying susceptibility genotypes [28]– [30], [46]. Interestingly, not all mutations in the *pncA* gene are linked to PZA resistance; some result in PZA-sensitive strains [45]. This complexity is illustrated in Figure 4. Consequently, assessing POA production in the culture medium offers a more comprehensive understanding of PZA susceptibility compared to isolated enzymatic activity measurements alone.

**Figure 4.**
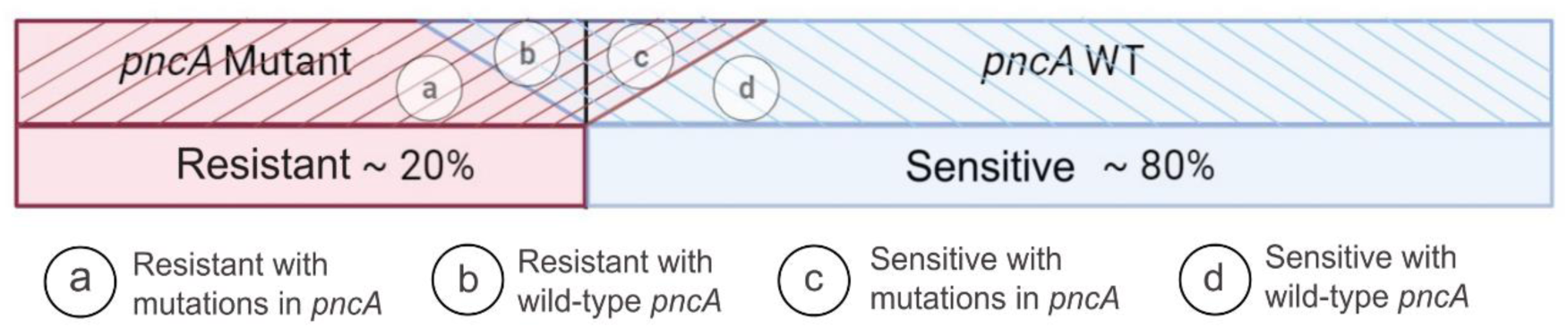
Complexity of PZA susceptibility phenotype. Variability in susceptibility phenotype is result of different factors such as, lack of specific PZA target, the large number of potential mutations throughout the coding and promoter sequence of *pncA*, the diversity of phenotypic methods and errors that they entail, among others. In this figure we can observe the 4 types of isolates according to the susceptibility phenotype.

In conclusion, the MODS-Wayne quantitative (MODS-WQ) assay in citrate buffer, with its exceptional predictive capacity (ROC: 97.6%, Kappa index: 0.87) and an average incubation period of just 13 days from sputum sample culturing, represents a significant advancement in TB diagnostics. This assay comprehensively accounts for factors like enzymatic efficiency and efflux pump activity, crucial for determining POA production in the medium. Additionally, its cost-effectiveness is particularly beneficial for resource-limited laboratories, especially in developing regions.

## Data Availability

All data produced in the present study are available upon reasonable request to the authors

## Acknowledgments

The authors extend their gratitude to the Hospital Nacional Dos de Mayo, Lima, Peru, and the Regional Tuberculosis Reference Laboratory, Callao, Lima, Peru for providing the sputum samples.

## Author contributions

Contributors ETG, MZ and PS conceived and designed the study. ETG analyzed data, generated figures and tables and wrote the manuscript. ETG, and RA collected clinical samples and data. ETG, and RA performed experiments, and analyzed data. PS, RG, LG and MZ contributed to data interpretation and critically reviewed the manuscript. All authors approved the final manuscript for submission.

**The authors declare no conflicts of interest.**

