## Supplementary Figures and Tables for "Quantitative MODS-Wayne Assay for Rapid Detection of Pyrazinamide Resistance in *Mycobacterium tuberculosis* from Sputum Samples"

**Toscano-Guerra E. *et al*.**

#### Supplemental information

**Figure S1. Schematic figure of for MODS, MODS-Wayne and MODS-WQ plates. A)** Layout of samples in the 24-well plate for MODS and MODS-Wayne assay, Control wells: growth control, RIF: rifampicin well, INH: isoniazid well, PZA: pyrazinamide well. M: sample. **B)** Layout of samples in the 24-well plate for two MODS-WQ variants (different plates). **C)** Representative coloration intensity in each sample. **D)** Schematic representation of the coloration intensity level according to the susceptibility.

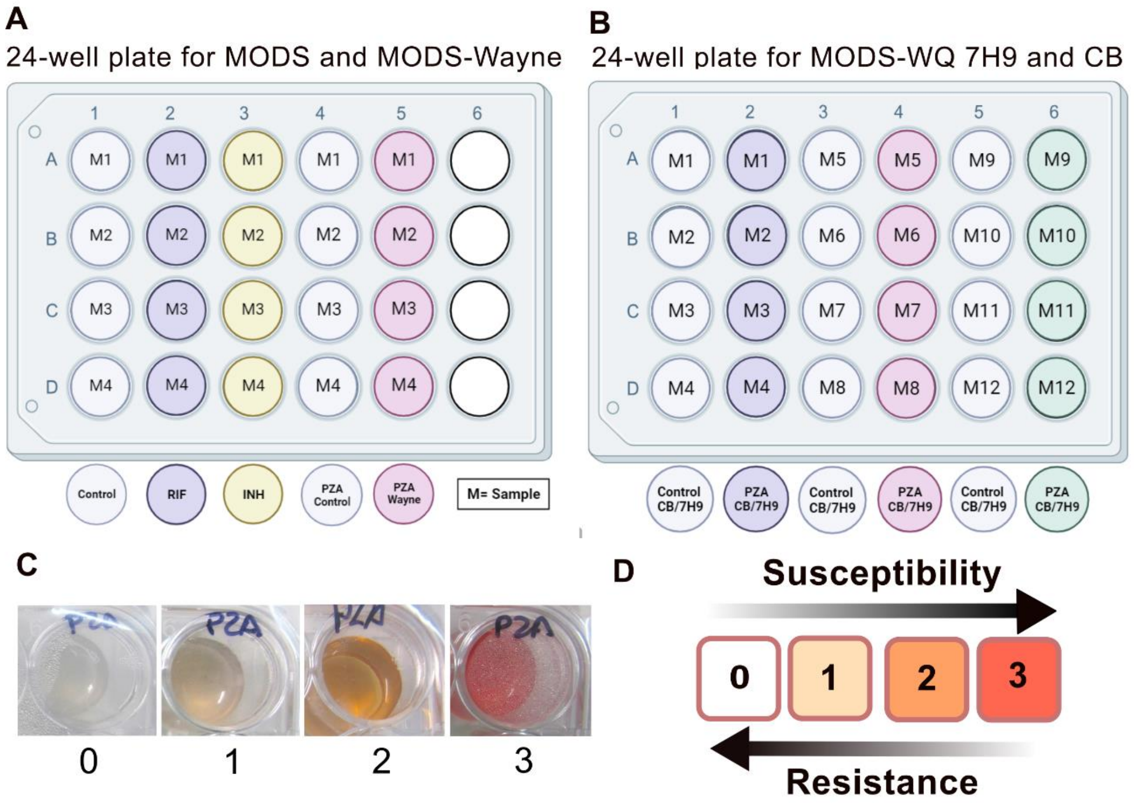

**Figure S2. Establishment of the composite standard. A)** Each isolate sample is cultivated in solid media, from which three reference methods were performed: BACTEC MGIT (a), Wayne test (b) and Sanger sequencing (c). **B)** Sample categorization according to references results, sensitive samples were coded as 0, resistant samples as 1. The sum of the results in each reference method is 0 or 1 for sensitive samples and 2 or 3 for resistant samples, according to the composite standard.

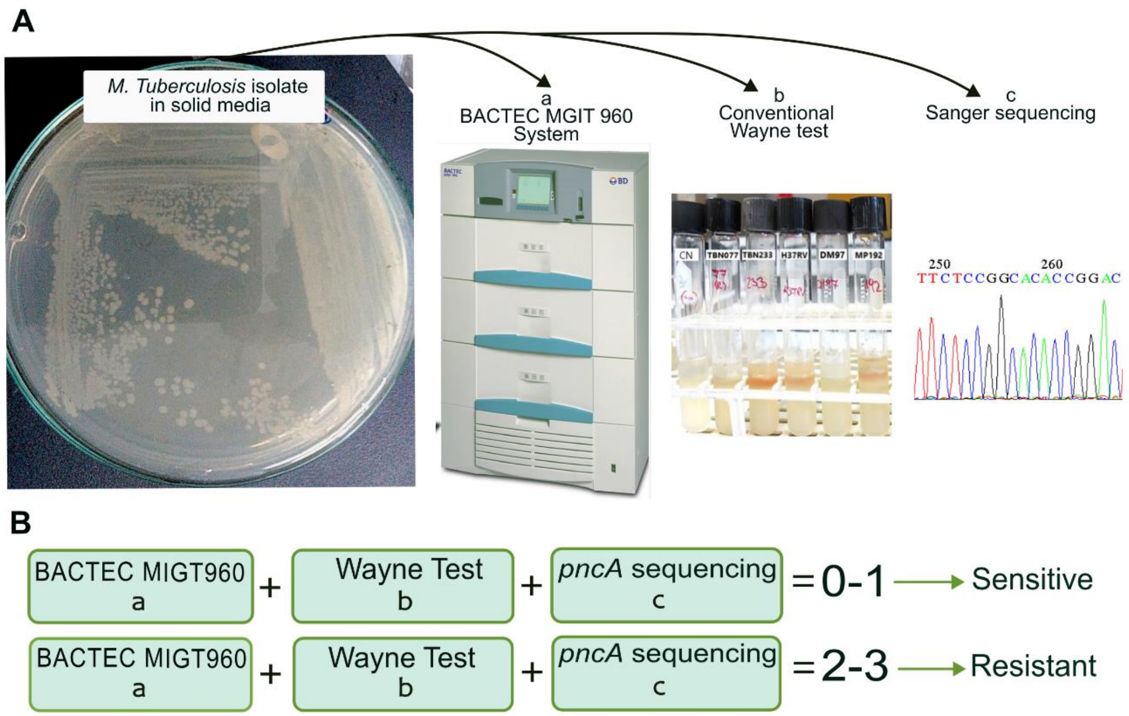

### TABLES

**Table S1. Summary of results with the three standard references**

| | MP (n=169) | | | TBN (n=95) | | |
|---|---|---|---|---|---|---|
| | **Wayne** | **MGIT960** | **Sequencing** | **Wayne** | **MGIT960** | **Sequencing** |
| **Sensitive** | 137 | 122 | 116 | 83 | 80 | 71 |
| **Resistant** | 32 | 47 | 48 | 12 | 15 | 22 |
| **Total** | 169 | 169 | 163* | 95 | 95 | 93* |

* Some samples were no able to be sequenced

**Table S2. POA concentrations for standard curve**

| **STOCK POA (mM)** | POA (µL) | Citrate Buffer /7H9-OADC (µL) | Final concentration POA (µM) | SAF 10% (µL) |
|---|---|---|---|---|
| **40** | 50 | 400 | 4000 | 50 |
| **20** | 50 | 400 | 2000 | 50 |
| **10** | 50 | 400 | 1000 | 50 |
| **5** | 50 | 400 | 500 | 50 |
| **2.5** | 50 | 400 | 250 | 50 |
| **1.25** | 50 | 400 | 125 | 50 |
| **0.625** | 50 | 400 | 62.5 | 50 |
| **0.312** | 50 | 400 | 31.25 | 50 |
| **0 (Agua)** | 50 | 400 | 0 | 50 |

**Table S3 . Summary of PZA susceptibility results according to 3 methodologies**

| Phenotype | Standard composite | | MODS-Wayne | | MODS-WQ | |
|---|---|---|---|---|---|---|
| | MP | 7H9 | MP | 7H9 | MP-BC | TBN-7H9 |
| Resistant | 46 | 16 | 41 | 15 | 43 | 31 |
| Sensitive | 123 | 79 | 123 | 80 | 121 | 64 |
