## Supplementary Methods for "Quantitative MODS-Wayne Assay for Rapid Detection of Pyrazinamide Resistance in *Mycobacterium tuberculosis* from Sputum Samples"

**Toscano-Guerra E.** ***et al.***

### Supplementary Methods

Sputum sample decontamination:

The samples were decontaminated according to the protocol by Kent and Kubica [31]. Each sample was mixed with a 2 % NaLc-NaOH solution in an equivolumetric ratio and incubated at room temperature for 15 min. After digestion with NaLc-NaOH, 10 mL of phosphate buffer pH 6.8 was added. It was homogenized by inversion and centrifuged (Heraeus X1R Centrifuge Thermo-Scientific, USA) at 3000 g for 15 min. Finally, the pellet is resuspended in 15 mL of 7H9 medium (BD Difco, USA) and stored at 4 °C until seeding.

Multidrug resistant test: rifampicin (RIF) and isoniazid (INH) resistance:

100 µl of 7H9 medium was dispensed in the first well on a 24-well plate (control well). Subsequently, 100 µl of drug solution (final concentrations, 0.4 µg/ml INH, 1.0 µg/ml RIF) was dispensed into the next two wells. Then 900 µl of decontaminated samples were added to each well. The plates were incubated at 37°C for a maximum of 21 days. The plate was read in an inverted microscope.

MODS-Wayne qualitative for PZA susceptibility:

Two wells per sample were used, the PZA control well (PZA-C) and the Wayne well (PZA-W). Then 900 µL of decontaminated samples was added to both wells and incubated at 37°C as in previous MODS assays. The PZA-C wells were evaluated every two dasy starting on the 5th day of incubation. Upon observing growth in the control well, the day was recorded. After three days, 100 µL of PZA was added to the PZA-W wells at a final concentration of 800 µg/mL. Then they kept incubating three more days with the PZA. At the end of the 3-day incubation with PZA, colorimetric reading of samples was conducted (only samples with positive growth in the control well were considered). For this, 100 µL of 10% ferrous ammonium sulfate (SAF) was added to both the Control-

PZA and Wayne-PZA wells, allowing a reaction time of 5 minutes, and the color was registered immediately. A sensitive result was reported when a red color was observed in the PZA-W well but not in the PZA-C well, and a resistant result was reported when no color was observed in both evaluated wells.

<u>MODS-Wayne quantitative susceptibility test:</u>

The MODS-WQ variants: MODS-WQ BC (10 mM citrate pH 7.0) and MODS-WQ 7H9 (pH ~6.8), each one in different 24-well plate. Samples (900 uL) were distributed in duplicates, the control well (PZA-C) and the drug well (PZA-WQ). Plates were incubated and evaluated as in MODS-Wayne assay. After the third day of PZA-C positive growth, samples were treated according to the variant. MODS-WQ 7H9 samples were completed to 1 mL with 100 µL of 7H9 media in PZA-C well and 100 µL of PZA (800 µg/mL final concentration) in the PZA-WQ well, and finally incubated for 3 additional days. Conversely, MODS-WQ CB samples were transferred to 2 mL tubes and centrifuged at 13000 rpm for 1 min. The resulting pellet was resuspended in 900 µL of citrate buffer and moved back to a 24-well plate. 100 µL of citrate buffer was then added to the PZA-C well and 100 µL of PZA (800 µg/mL final concentration) to the PZA-WQ well. It was then incubated for an additional 3 days. After incubation, 100 uL of 10% SAF was added and incubated for 5 minutes. Then 500 µL were transferred to cryotubes and stored at - 80°C, until its subsequent reading. In the reading day, samples were transferred (100 uL) in triplicates into a 96-well plate and were read at 450nm wavelength on a spectrophotometer. Of note, MODS-WQ 7H9 samples were previously centrifugated to 13000 rpm for 2 minutes The absorbance values were interpolated to the standard POA curves, according to the matrix variant.

<u>Calibration curve for pyrazinoic acid:</u>

For calibration curves for POA (Sigma-Aldrich P56100) was used at 8 final concentrations, from 31.25 µM to 4000 µM. Calibration curves with 7H9 matrix were previously centrifugated at 13000 rpm for 2 minutes and supernatants were measured. The mixtures (100 uL) were transferred to a 96-well plate in triplicates to be read at 450nm wavelength on a spectrophotometer. The absorbance values were used to the calculate the lineal regression of standard curves.

<u>Composite reference standard for PZA susceptibility determination:</u>

To perform the standard tests, a pure isolate of each sample was required. In brief, *M. tuberculosis* was isolated from 100 µL of the control wells of the MODS-WQ test, on 7H10 (Middlebrook agar) enriched with OADC and incubated at 37 ° C for 21 to 28 days.

*pncA* gene sequencing

One full loop of *M. tuberculosis* culture was suspended in 500 µl of Tris-EDTA buffer at pH 8.0 and inactivated by heating to 80°C for 30 min. DNA extraction was performed using a modified proteinase K-chloroform protocol [33] and amplification of *pncA* gene was performed as previously described [30] using primer P1 (5′-GTCGGTCATGTTCGCATCG-3′; from 105 bp upstream of *pncA*) and primer P6 (5′-GCTTTGCGGCGAGCGCTCCCA-3′; from 60 bp downstream of *pncA*) [34]. The amplification product (720 bp) was sequenced using the same primers and the presence of mutations in *pncA* gene and putative promoter was evaluate by pairwise alignment using the wild type sequence of *M. tuberculosis* H37Rv reference strain (NC_000962.3). Identified mutations were compared with the Catalogue of mutations *in Mycobacterium tuberculosis* [35] developed by WHO and the *pncA* mutation tool SUSPECT-PZA [36].

Conventional Wayne test

One heavy loopful was inoculated onto Dubos culture medium. Tubes were incubated at 37°C for 7 days. After this, 1 mL of 1% SAF was added to reveal the production of POA. The reaction was incubated for 30 minutes at room temperature. The formation of a pink ring on the surface of the medium indicated positive production of POA and the isolate was registered as PZA-sensitive. Tubes without a ring were further incubated at 4°C for 3 hours, If no ring formed it was considered negative and the isolate was registered as PZA-resistant [18]. For each test round, positive (H37Rv) and negative (DM097) controls were included.

BACTEC MGIT 960

The MGIT-PZA was performed in accordance with manufacturer's instructions. In brief, a suspension with turbidity equal to 0.5 McFarland was prepared. The suspension was diluted in physiological saline at a ratio of 1:5 and 1:10, 0.5 ml of which was added to the susceptibility test tube and control tube, respectively. In addition, PZA was added to the susceptibility test tube at a final concentration of 100 µg/ml. The tubes were incubated in a MGIT 960 system.
